# Seroprevalence of anti-SARS-CoV-2 antibodies in Iquitos, Loreto, Peru

**DOI:** 10.1101/2021.01.17.21249913

**Authors:** Carlos Álvarez-Antonio, Graciela Meza-Sánchez, Carlos Calampa, Wilma Casanova, Cristiam Carey, Freddy Alava, Hugo Rodríguez-Ferrucci, Antonio M. Quispe

**Affiliations:** Dirección Regional de Salud de Loreto, DIRESA, Loreto, Peru; Universidad Nacional de la Amazonía Peruana, Loreto, Peru; Centro de Investigación en Bioingeniería, Universidad de Ingeniería y Tecnología, Lima, Peru; Universidad Continental, Huancayo, Peru

**Keywords:** Severe Acute Respiratory Syndrome, Coronavirus 2, COVID-19, Seroepidemiologic Studies, Peru

## Abstract

**Background:** Detection of SARS-CoV-2 antibodies among people at risk is critical for understanding both the prior transmission of COVID-19 and vulnerability of the population to the continuing transmission and, when done serially, the intensity of ongoing transmission over an interval in a community. In this study, we estimated the seroprevalence of COVID-19 in a representative population-based cohort of Iquitos, one of the regions with the highest mortality rates from COVID-19 in Peru, where a devastating number of cases occurred in March 2020.

**Methods:** We conducted a population-based study of transmission tested each participant using the COVID-19 IgG/IgM Rapid Test from Orient Gene Biotech and used survey analysis methods to estimate seroprevalence accounting for the sampling design effect and test performance characteristics. Here we report results from the baseline (13 to 18 July 2020) and the first month of follow-up (13 to 18 August 2020) study.

**Findings:** We enrolled a total of 716 participants and estimated seroprevalence of 70.0% (95% CI: 67.0%–73.4%), a test-re-test positivity of 65% (95% CI: 61.0%–68.3%), and an incidence of new exposures of 1.8% (95% CI: 0.9%–3.2%) data that suggest that transmission is ongoing but is occurring at low levels. We observed significant differences in the seroprevalence between age groups, with participants 18 to 29 years of age having lower seroprevalence than children <12 years of age (Prevalence ratio =0.85 [PR]; 95% CI: 0.73 – 0.98), suggesting that children were not refractory to infection in this setting.

**Interpretation:** Iquitos demonstrates one of the highest rates of seroprevalence of COVID-19 worldwide. Current data shows a limited case burden in Iquitos for the past seven months and suggests that these levels are sufficient to provide significant but incomplete herd immunity.

**Funding:** Dirección Regional de Salud de Loreto, DIRESA, Loreto, Peru

## Introduction

COVID-19 was first recognized in Peru in March 2020^1^, causing more than 90,000 deaths and over one million confirmed infected by August 15, 2020, with presumably millions more infected.^2^ One component of an effective Public Health response to the pandemic is determining what proportion of the population remains susceptible and immune to the virus and assessing the duration of protection; these questions are best answered through seroprevalence surveys.^3^ The presence of SARS-CoV-2 antibodies indicates prior exposure^4^ and has shown with very few exceptions that these individuals are protected against reinfection.^5^

SARS-CoV-2 infections are either symptomatic, pre-symptomatic, or asymptomatic, with asymptomatic individuals having comparable viral loads to symptomatic cases^6^ and playing an essential role in transmitting the disease.^7^ Without symptoms, most asymptomatic cases are never being noticed, mainly in seroprevalence studies. Nevertheless, seroprevalence estimates vary widely depending on country and risk groups. For example, some states such as San Francisco in the United States reported seroprevalences as low as 0.26%^8^; Wuhan China, 3.2-3.8%^9^; while Switzerland, 11%^10^; New York City, 19%^11^, and more recently the highest reported in Manaus, Brazil, with 55.1-61.4%.^12^ In Peru, preliminary results from two seroprevalence studies, one carried out in July, reported a seroprevalence of 29.7% in the region of Lambayeque. Another one has carried out in December, a seroprevalence of 39.3% in Lima and Callao’s regions using population-based sampling techniques.^13^

Herein we report SARS-CoV-2 seroprevalence rates from Iquitos, Peru, one of the first and hardest-hit cities during the pandemic in Peru. In Iquitos city, over two-thirds of the total excess of deaths occurred over one month (April 19-May18), including the death of 23 physicians and the health care system collapse.^14^ The study was carried out by the Directorate of Epidemiology, Executive Directorate of Prevention and Control of Diseases, Regional Directorate of Health of Loreto, from July-August 2020, following a Public Health Emergency in April and May 2020 when over one thousand Iquitos residents, including over 50 health workers, died during the first wave of the Pandemic.

This study informs decision-making aimed at mitigating the further impact of COVID-19.

## METHODS

### Ethics Statement

The study was approved by the hospital health network Institutional Scientific Committee and Institutional Review Board of the Regional Hospital, Iquitos. Written informed consent was obtained from all adult participants and parents or a legal representative of all children < 18 years of age. Also, written assent was provided by children ≥13 to <18 years of age. Participants’ data were coded to protect their identity. Study forms and codes were protected and handled only by the study researchers.

### Study Setting

Iquitos (73.2°W, 3.7°S) is a city of ∼467,000 inhabitants, located in the department of Loreto in northeast Peru. The city has four districts, San Juan to the south, Belen to the East, Punchana to the North, and Iquitos in the city’s central part.^15^ As most Peruvian Amazon cities, Iquitos is accessible from the coast only by air or by boat, situated at 120 meters above the sea level at the confluence of the Amazon, Itaya, and Nanay Rivers. The climate is tropical rainforest, with daily temperatures of 22.0–32.2°C, with the coolest months observed typically in June and July (21.0–31.1°C), and the warmest months in October through December (26·2°C [22·5–33·1°C]). The average annual precipitation is 3.6 meters, with the rainiest months typically observed from December through May. According to the latest census, Iquitos ranks among Peru’s poorest regions, with more than 60% of Iquitos population classified either as poor or extremely poor. The regions’ principal industries are oil, tourism, agriculture, fishing, and lumber. Iquitos sanitary infrastructure includes one military clinic, one military hospital, three public hospitals, and nine public clinics, most of them located in Iquitos’ urban areas. The Peruvian government is currently providing full coverage for COVID-19 healthcare, so these patients receive medical attention free of charge. The regions have an autochthonous transmission of all four distinct dengue virus serotypes since 1990, with serotype two currently being the most predominant^15^. Yellow fever is endemic in the region^16^, and the Zika virus was introduced into Iquitos in late 2016^17^.

### Sample size

The minimum sample size was determined for an underlying crude SARS-CoV-2 seroprevalence of 18% or higher during the study period. Sampling effects were calculated to weight for the district population and population distribution by gender and age groups (< 12 years, 12 to 17 years, 18 to 29 years, 30 to 59 years, and > 60 years). The effective sample size was approximately 400 subjects to estimate a seroprevalence of 18% with a precision of ±2.5% (50% relative error) at the 95% confidence level. In anticipation of a 20% response rate, 20% loss in follow-up, and 20% missing values, the minimum sample to be selected increased to 692 subjects. Based on this estimate, we decide to enroll 726 participants to account for a 5% loss of information due to contingencies like robberies, assaults, or similar.

### Study design

We carried out a population-based 3-month cohort study between July-August 2020 of a geographically stratified sample across Iquitos city. We obtained a representative sample of the Iquitos city population using the 2017 census data, which the Ministry of Health updated on January 20, 2020, for vaccination purposes. This information includes detailed maps with the limits of each of the four levels considering in our sampling procedure, including four districts, 40 sectors, 2500 blocks of households, and 90,354 households. We aimed to obtain a representative sample of each district, age group, and gender, so we weight the sample by districts, gender, and age groups. At each district, we sample each sector, and at each selected random house, we search for an individual of a specific age group and gender. If participants were not available, we replaced the house with nearby households from right to left until we find a match. In all cases, eligible participants were invited to participate, and all those who gave their informed consent were surveyed. We attempted to screen each of the cohort participants during the first week of each month for three consecutive months, July, August, and September. Additionally, we reported the regional excess death counts as reported by the National Death Registry Information System (SINADEF) to contextualize de Study.

### Inclusion and Exclusion Criteria

We included in our study all the inhabitants of Iquitos, Loreto, Peru, since the arrival of COVID-19 to Peru (March 6^th^, 2020). Exclusion criteria included: (a) institutionalized individuals at nursing homes, prisons, or boarding schools, (b) subjects receiving any pharmacological treatment proposed for COVID-19 (hydroxychloroquine, ivermectin, or azithromycin), (c) individuals with any contraindication for phlebotomy (cellulitis or abscess, venous fibrosis on palpation, presence of hematoma, vascular shunt or graft, or a vascular access device), and (d) health workers or individuals living with an active health worker.

### Procedures

Study personal visited each eligible subject and asked participants for their voluntary participation. After providing consent, the study personal interviewed the participant using standardized ministry of health epidemiological investigation forms. To detect IgG and IgM anti-SARS-CoV-2 antibodies, we used COVID-19 IgG/IgM Rapid Test Cassette rapid diagnostic test from Zhejiang Orient Gene (Biotech Co LTD, China) on capillary blood obtained by finger-prick. Test results were provided to each participant on-site with instructions about how to proceed if tested negative or positive.

### Laboratory analysis

The COVID-19 IgG/IgM Rapid Test Cassette, a rapid diagnostic test from Zhejiang Orient Gene (Biotech Co LTD, China), is an immunochromatographic assay uses SARS-COV-2 antigen-coated particles to qualitatively detect IgG and IgM antibodies in whole blood, capillary blood, serum, and plasma. Following the manufacturers’ instructions, we waited 10 minutes to read the test results and verified that the first band (control) was observed, indicating that the test performed properly. This test was independently evaluated by Delliere et al^18^, who reported a sensitivity of 95.8% (95% confidence interval [CI95%], 89.6% to 98.8%) and a specificity of 100% (CI95%, 93.4% to 100%) for COVID-19 seroprevalence. The first read was performed by the study personal on-site. In contrast, the second and, if needed, the third read was performed centrally using high-quality digital images of the test result immediately took on site after the first read. We performed a quality control double read and use a third independent interpretation to resolve any disagreement between the first two readers.

### Statistical analysis

We performed a descriptive analysis summarizing the participant’s demographics and clinical history with absolute and relative frequencies if categorical and with their mean and standard deviation if continuous. We use Student’s t-test for mean comparisons and Fisher’s exact test for proportion comparisons between those that tested seropositive versus those that tested seronegative. Then, we estimated the seroprevalence using a two-step process. First, we estimate the seroprevalence by accounting for the survey sampling weights using STATA survey (svy) commands and, second, we adjusted these estimates to account for the sensitivity (95.8%) and specificity (100%) using the diagnostic (diagti) command. Finally, we explored associated factors to three outcomes of interest, including COVID-19 seropositivity (defined as positive at baseline), and test-re-rest positivity (defined as positive at baseline and positive at the first month follow up), and incidence of new COVID-19 exposure (defined as negative at baseline and positive at the first month follow up). In all cases, we estimated the prevalence ratio as the magnitude of association of interest using log-binomial regression models with robust variance and a confidence interval of 95%. We performed all the data analysis using the statistical package STATA^tm^ MP version 14.0 (Stata Corp LP, College Station, Texas).

## Results

### Study population

At baseline, we enrolled a total of 716 out of 726 eligible subjects, which were distributed in 40 strata (four districts, two genders, and five age groups). We excluded ten eligible subjects because some did not consent their children to participate (n =3), some recently moved to Iquitos (n =3), others were subjects in transit (n =2), or one had respiratory symptoms. We referred this patient to the regional hospital, where he was further evaluated and tested negative for COVID-19.

### *Population* characteristics

We included a representative sample from each of the Iquitos four districts (Table 1). Participant’s mean age was 29.2 years of age (range: 3 months to 89 years of age). Most patients were women (51.0%), from San Juan (42.0%) or Iquitos (36.6%) districts, and proceed from urban areas (86.5%). Most patients denied any history of previous existent conditions (85.6%), whereas the following were reported: cardiovascular diseases (6.0%), diabetes (2.8%), chronic respiratory diseases (2.0%), obesity (1.3%), and kidney disease (0.9%). Likewise, the study sample included four pregnant women and one subject with Down’s Syndrome.

**Table 1:** Characteristics of the study population

| Characteristic | Baseline (n = 716) |  | Follow-up (n =621) |  |
| --- | --- | --- | --- | --- |
|  | Positivity<br>N (%) | Seroprevalence<sup>1</sup><br>% (95% CI) | Positivity<br>N (%) | Seroprevalence<sup>1</sup><br>% (95% CI) |
| Test results |  |  |  |  |
| IgG+ or IgM+ | 528 (73.7) | 70.0 (67.0–73.4) | 422 (68.0) | 66.0 (62.0–70.1) |
| Total IgG+ | 526 (73.5) | 70.0 (66.0–73.0) | 379 (61.0) | 59.0 (55.0–63.3) |
| Total IgM+ | 50 (7.0) | 7.5 (5.7–9.7) | 43 (6.9) | 7.1 (5.2–9.4) |
| Only IgG+ | 478 (66.8) | 63.0 (59.0–66.1) | 379 (61.0) | 59.0 (55.0–63.3) |
| Only IgM+ | 2 (0.3) | 0.4 (0.1–1.2) | 5 (0.8) | 1.0 (0.4–2.2) |
| District |  |  |  |  |
| Belen | 88 (78.6) | 76.0 (67.0–83.5) | 61 (61.0) | 61.0 (51.0–70.6) |
| Iquitos | 173 (74.3) | 70.0 (64.0–75.8) | 149 (70.3) | 68.0 (61.0–74.2) |
| Punchana | 96 (73.3) | 71.0 (62.0–78.6) | 71 (67.6) | 67.0 (57.0–75.6) |
| San Juan | 171 (71.3) | 69.0 (62.0–74.6) | 141 (69.1) | 66.0 (59.0–72.6) |
| Gender |  |  |  |  |
| Women | 273 (72.4) | 70.0 (64.0–74.2) | 225 (68.6) | 66.0 (61.0–70.6) |
| Men | 255 (72.4) | 71.0 (66.0–75.4) | 197 (67.2) | 66.0 (60.0–70.6) |
| Age, years |  |  |  |  |
| < 12 years | 141 (76.2) | 70.0 (63.0–77.0) | 121 (71.2) | 69.0 (61.0–75.7) |
| 12 to 17 years | 70 (81.4) | 72.0 (62.0–81.5) | 51 (71.8) | 73.0 (61.0–83.1) |
| 18 to 29 years | 90 (64.8) | 54.0 (45.0–62.0) | 58 (50.9) | 50.0 (40.0–59.5) |
| 30 to 59 years | 186 (73.8) | 66.0 (60.0–72.1) | 152 (71.0) | 68.0 (62.0–74.4) |
| > 60 years | 41 (75.9) | 74.0 (60.0–85.0) | 40 (76.9) | 77.0 (63.0–87.5) |
| Rural/Urban |  |  |  |  |
| Rural | 55 (73.3) | 71.0 (59.0–80.6) | 48 (81.4) | 73.0 (60.0–83.6) |
| Urban | 473 (73.8) | 70.0 (66.0–73.6) | 381 (67.8) | 65.0 (61.0–69.4) |

<sup>1</sup> Seroprevalence estimated considering sample effects and independent estimate of the sensitivity (95.8%) and specificity (100%) of the diagnostic test for seroprevalence studies.

### Seroprevalence estimate

At baseline, we observed seropositivity of 73.7% (528/716) either to IgM (7.0%) or IgG (73.5) anti-SARS-CoV-2 antibodies. After adjusting for the study sampling effects and the independently reported sensitivity (95.8%) and specificity (100%) of the rapid test, we estimated seroprevalence of 70.0% (95% CI: 67.0% to 73.4%) at baseline. At the first month follow-up, we observed a positivity of 68.0% (422/621) either to IgM (7.5%) or IgG (70.0%) anti-SARS-CoV-2 antibodies. After accounting for the study sampling effects and the test sensitivity and specificity, we estimated seroprevalence of 66.0% (95% CI: 62.0% to 70.1%). Here is important to highlight that the study baseline was carried out two months after the peak of the epidemic curve in Iquitos (Figure 1).

**Figure 1:**
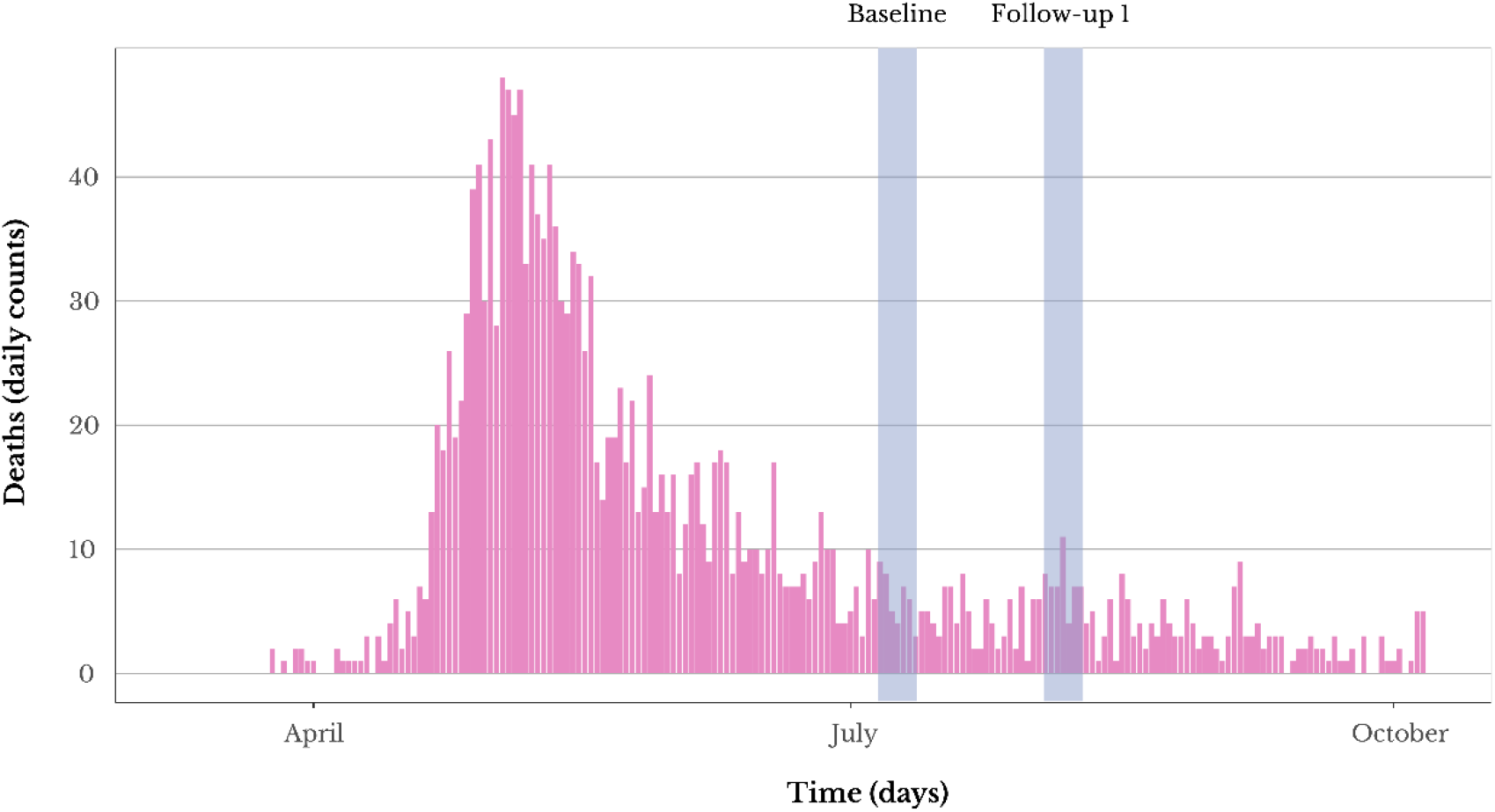
Daily excess of deaths at Iquitos city as reported by the SINADEF. Blue squares represent the periods of time during which we carry out the study baseline and follow-up 1. Deaths counts only includes those reported as non-violent deaths due any cause, which during the pandemic were reported as the strongest and most reliable signal to follow-up the track to the COVID-19 epidemic in Peru.

### Seroprevalence variability

Overall, we observed that the seroprevalence in Iquitos varied significantly across age groups. Specifically, we observed that the seroprevalence was the highest among participants from extreme age groups, meaning < 12 years (70.0%; CI 95%: 63.0% to 77.0%) and ≥ 60 years (74.0%; CI 95%: 60.0% to 85.0%), and the lowest among participants with age 18 to 29 years old (54.0%; CI 95%: 45.0% to 60.0%). We did not observe significant differences by districts, gender, and urban/rural areas.

### Test-re-test and incidence analysis

We performed a test-retest analysis in the 621 (86.7%) participants that completed the first month of follow-up (Figure 2). Among them and after adjusting for the study sampling effects and the test’s sensitivity and specificity, we estimated a COVID-19 test-re-test positivity of 65% (95% CI: 61.0% to 68.3%). Likewise, we estimated an incidence of new COVID-19 exposures of 1.8% (95% CI: 0.9% to 3.2%).

**Figure 2:**
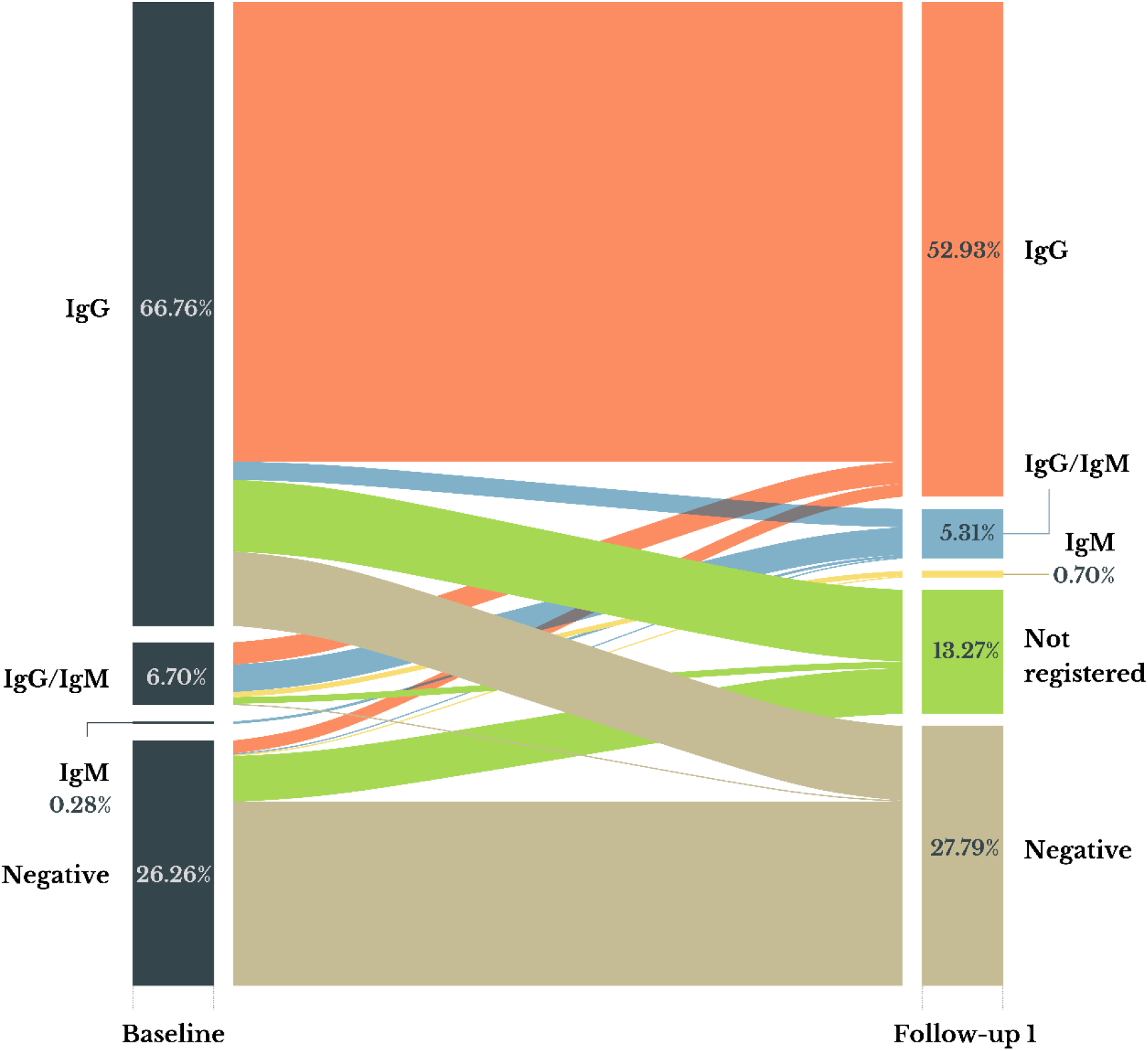
Distribution of the study participants based on their IgM and IgG antibodies anti SARS-CoV-2 at baseline and follow-up 1. Notice that we loss 12.27% of the study population at the follow-up 1, which are reported here as “not registered. Notice that even when both, baseline and follow-up 1 have a similar seropositivity (73.7% vs. 68%), a significant fraction of the participants that tested positive for IgG at baseline tested negative at the follow-up 1 (∼28%).

**Table 2:** Factors associated with the seroprevalence, test-re-test positivity and incidence of IgM or IgG anti-SARS-CoV-2 antibodies of COVID-19 in Iquitos City

| Factor | *Seroprevalence*<br>PR (CI 95%) | *Test-Re-Test*<br>PR (CI 95%) | *Incidence*<br>PR (CI 95%) |
| --- | --- | --- | --- |
| Age, years |  |  |  |
| < 12 years | Ref. | Ref. | Ref. |
| 12 to 17 years | 1.07 (0.94–1.22) | 1.00 (0.83–1.21) | 1.20 (0.22–6.40) |
| 18 to 29 years | 0.85 (0.73–0.98)<sup>†</sup> | 0.71 (0.58–0.88)<sup>‡</sup> | 0.75 (0.14–4.01) |
| 30 to 59 years | 0.97 (0.87–1.08) | 1.01 (0.88–1.16) | 0.60 (0.14–2.63) |
| > 60 years | 0.99 (0.84–1.18) | 1.09 (0.90–1.31) | 0.82 (0.09–7.16) |
| Rural/Urban |  |  |  |
| Rural | Ref. | Ref. | Ref. |
| Urban | 1.01 (0.77–1.09) | 0.92 (0.77–1.09) | 0.21 (0.07–0.68)<sup>‡</sup> |
| Gender |  |  |  |
| Female | Ref. | Ref. | Ref. |
| Male | 0.97 (0.88–1.05) | 0.98 (0.87–1.09) | 1.12 (0.36–3.44) |
| District |  |  |  |
| Belen | Ref. | Ref. | Ref. |
| Iquitos | 0.94 (0.84–1.07) | 1.19 (0.98–1.44) | 0.47 (0.10–2.30) |
| Punchana | 0.93 (0.81–1.07) | 1.15 (0.93–1.42) | 0.32 (0.03–3.01) |
| San Juan | 0.91 (0.80–1.03) | 1.15 (0.95–1.36) | 0.82 (0.20–3.35) |

\* PR, prevalence ratio; <sup>†</sup>, p-value <0.05; <sup>‡</sup>, p-value <0.01

### Associated factors regression analysis

At baseline, we observed that the COVID-19 seroprevalence was associated with age, with subjects 18 to 29 years of age having significant lower COVID-19 seroprevalence than subjects <12 years of age (Prevalence ratio =0.85 [PR]; 95% CI: 0.73 to 0.98; *p*-value =0.029). Similarly, we observed that among those that completed the first-month follow-up, the rest-re-test seropositivity was associated with age, with subjects 18 to 29 years of age having significant lower COVID-19 seroprevalence than subjects <12 years of age (PR=0.71; 95% CI: 0.58 to 0.88; *p*-value =0.002). Additionally, we observed that among those that completed the first month of follow-up, the incidence of new COVID-19 exposures was associated with the origin of the participant being lower if the proceed from an urban area compared to from a rural area (PR=0.21; 95% CI: 0.07 to 0.68; *p*-value =0.009). We attempt to progress to multivariable regression analysis, but AICs suggested that we keep the first-order models at each regression.

## Discussion

The findings from this population-cohort study for SARS-CoV-2 indicate that IgG/IgM antibodies’ seroprevalence against this coronavirus is around 70% at Iquitos. Because the study was designed to obtain a representative sample from the Iquitos inhabitants, we observed differences across age groups that are consistent with the literature. Moreover, it is noticeable that we observed significantly higher seroprevalence among children under 12 years of age than the seroprevalence observed among adults 30 to 59 years of age, despite school closure and strictly mandated restrictions in social gatherings during the study period. Furthermore, we observed a seroprevalence of COVID-19 among Iquitos children was as high as the one observed among adults over 60 years of age.

To our knowledge, our study describes one of the highest seroprevalences of COVID-19 in the world. Previously, high seroprevalence estimates were reported in Mumbai, India, and Manaus, Brazil. In Mumbai, between June 29 and July 19, 2020, local researchers reported a seroprevalence between 55.1% and 61.4% using a population-based sample.^12^ While in Manaus, using blood donors samples, researchers reported in July 2020 a seroprevalence of SARS-CoV-2 between 44% and 66%.^19^ With a population of around 426,000, the city of Iquitos was one of Peru’s cities that was hit the harder by the COVID-19 pandemic. At the peak of Iquitos COVID-19 epidemic, in the first week of May, the regional authorities declared a major sanitary crisis with over 100 deaths per day, which added up to around 2500 registered deaths either confirmed (∼1200) or suspected (∼1300) and many more deaths due to natives unregistered burials. Despite government efforts, hospitals became overwhelmed, and medical oxygen shortages contributed mainly to these deaths. The tragic news reports gave us a sense of the magnitude of the humanitarian crisis in Iquitos. Known as the world’s largest city that cannot be reached by road, Iquitos suffered a COVID-19 epidemic of a catastrophic proportion, despite imprecisions in exact case counts and incomplete deaths attribution. Our study estimates show that the vast majority of Iquitos inhabitants were infected with COVID-19 before our baseline measure. However, the transmission did appear to continue at a low level, as seen by a slight increase in seroprevalence in the test-retest findings.

We found no differences in seroprevalence between females and males among districts or origin from rural and urban areas. In contrast to what has been reported previously, we observed the highest SARS-CoV-2 seroprevalence among children, teenagers, and elders, and the lower SARS-CoV-2 seroprevalence among adults throughout the age 18 to 59 years of age. This observation is counterintuitive given that children in most similar studies resulted with the lower prevalence presenting the to lower nasal gene expression of angiotensin-converting enzyme 2.^20^ A possible explanation for these findings is that Iquitos is one of the poorest cities in Peru, most citizens lack potable water and sanitation, and only a small fraction poses the means to preserve their food refrigerated. This means that most families need to trade or buy food daily, and these needs exposed children to the community to a more considerable extent than in different social contexts. This is particularly relevant because, in mid-May, the Peruvian Ministry of Health conducted a series of surveillance studies on the country’s largest markets using the same test used in our research, finding a seroprevalence among the sellers at the Belen market of 99.9%.

Seroprevalence studies are essential to estimate the number of COVID-19 infections, mainly because of the high rate of asymptomatic and pre-symptomatic that often undergo testing.^21^ In an example, the USA reported that the estimated number of total COVID-19 infections ranged between 6 to 24 times the number of confirmed cases, after accounting for the asymptomatic/pre-symptomatic individuals that undergo testing.^22^ Furthermore, seroprevalence studies are becoming essential to assess the risk of second and third waves of COVID-19 in all countries. For example, in mid-August, it was estimated that the pooled seroprevalence of SARS-CoV-2 antibodies at a global level was 3.38% (IC95% 3.05-3.72), with Latin America having just 1.45% (IC95% 0.95-1.94).^23^ This means that the risk of a second wave and a third one, in the absence of a vaccine, remained high and gave the marked variability of SARS-CoV-2 seroprevalence among and within each geographic region; such waves also will vary substantially.

Our study’s key strength is the random selection of households from the Iquitos census (updated on Jan 20, 2020), which allowed us to obtain a representative sample of the inhabitants of Iquitos city. Furthermore, the study was overpowered because the high seroprevalence was four times higher than expected. Even though we lost 13% of participants at the first follow-up, all four districts were adequately represented in our study. Participation rates were slightly lower in rural areas, but this was compensated by the high seroprevalence observed across Iquitos regions. Our analysis only detected IgM and IgG antibodies, but the extent of the immunity they provide is uncertain, with cellular immunity also playing a role in protecting against SARS-CoV-2 reinfection.^24^

During the first wave of COVID-19, Latin America become the epicenter of the pandemic with Brazil and Peru as the worst-case scenarios, despite their different control approaches. While Brazil mostly decided to pursue herd immunity, Peru implemented one of the longest and strict lockdowns early in the pandemic. However, Iquitos was one of the last cities in Peru to enforce the lockdown policies and experienced one of the most tragic COVID-19 epidemics globally. In less than four weeks, Iquitos COVID-19 epidemic peak increasing 6-8 times in cases and 4-5 times in deaths reaching the higher mortality in Peru at the time. Currently, mortality rates in Iquitos have returned to baseline, which provides support for our findings.

Iquitos appears to have become one of the first cities worldwide to surpass the herd immunity threshold, which has been estimated for COVID-19 to be 60-70%^25^. Although it is clear that also Iquitos paid a huge cost in terms of human suffering and deaths, our findings appear to support model-derived estimates for the level of seropositivity needed to provide herd immunity.

## Data Availability

The authors confirm that the data supporting the findings of this study are available within the article.

## Contributors

CAA, GMS, WC, HRF, CrC, and AMQ were responsible for the study’s conceptualization and design. CaC and GMS are the executive coordinators of the project and led the relationship with the DIRESA Loreto. CAA, GMS, CaC, CrC, and FA were responsible for obtaining the serological tests’ funding, the field personal, and producing the laboratory results. CAA, GMS, and FA are responsible for the study operation, including data acquisition and logistics coordination. FA and AMQ developed the operational protocols for fieldwork and were in charge of verifying the underlying data. FA was responsible for training the involved administrative and health personnel. AMQ was in charge of statistical analyses and table and figure design. CAA and AMQ wrote the first draft. All authors contributed to data interpretation, critically reviewed the first draft, approved the final version, and agreed to be accountable for the work.

## Data sharing

The anonymized, aggregated data collected to support this pooled data analysis is available in supplementary file 2 for non-commercial use only.

## Acknowledgments

This work was supported by the Loreto Health Directorate (DIRESA) and the Peruvian Ministry of Health. The DIRESA Loreto also provided the provided census information necessary for the random selection of households. We thank all the health professionals, administrative personnel, and other health-care workers who collaborated in this study and all participants. The authors thank the study participants and the DIRESA team from Loreto. Additionally, we would like to thank Enrique Mendoza for his support elaborating the graphics of the study and Valerie Paz Soldan, Amy Morrison, and Margaret Kosek for their review and feedback on the writing of the manuscript. This study is the result of the efforts of researchers from Loreto, all of which also suffered from COVID-19 during the Iquitos epidemic, and the trust and generosity of more than 700 participants who have understood the importance of providing time, information, and samples to learn about the COVID-19 epidemic in Iquitos.

## Disclaimer

The findings and conclusions presented in this report are those of the authors and do not necessarily reflect the Peruvian Ministry of Health’s official position.

## Supplementary Files

**Supplementary File 1:** Excel database of study data, including all variables and data dictionary.

